# Fine mapping of the *HLA* locus in Parkinson’s disease in Europeans

**DOI:** 10.1101/2020.10.29.20217059

**Authors:** Eric Yu, Aditya Ambati, Maren Stolp Andersen, Lynne Krohn, Mehrdad A. Estiar, Prabhjyot Saini, Konstantin Senkevich, Yuri L. Sosero, Ashwin Ashok Kumar Sreelatha, Jennifer A. Ruskey, Farnaz Asayesh, Dan Spiegelman, Mathias Toft, Marte K. Viken, Manu Sharma, Cornelis Blauwendraat, Lasse Pihlstrøm, Emmanuel Mignot, Ziv Gan-Or

**Affiliations:** Department of Human Genetics, McGill University, Montréal, QC, Canada; The Neuro (Montreal Neurological Institute-Hospital), McGill University, Montreal, QC, Canada; Stanford Center For Sleep Sciences and Medicine, Department of Psychiatry and Behavioral Sciences, Stanford University, Palo Alto, California, United States of America; Department of Neurology, Oslo University Hospital, Oslo, Norway; Institute of Clinical Medicine, University of Oslo, Oslo, Norway; Department of Neurology and Neurosurgery, McGill University, Montréal, QC, Canada; Centre for Genetic Epidemiology, Institute for Clinical Epidemiology and Applied Biometry, University of Tübingen, Tübingen, Germany; Department of Medical Genetics, University of Oslo and Oslo University Hospital, Oslo, Norway; Department of Immunology, Oslo University Hospital, Oslo, Norway; Molecular Genetics Section, Laboratory of Neurogenetics, National Institute on Aging, National Institutes of Health, Bethesda, MD, United States of America

**Keywords:** Parkinson’s disease, HLA, Genetic analysis

## Abstract

**Objective:** To fine map the association between human leukocyte antigen (*HLA)* genes and Parkinson’s disease (PD) that was discovered using genome-wide association studies (GWASs).

**Methods:** We performed a thorough analysis of the *HLA* locus in 13,770 PD patients, 20,214 proxy-cases and 490,861 controls of European origin. We used GWAS data to impute *HLA* types and performed multiple regression models to examine the association of specific *HLA* types, different haplotypes and specific amino acid changes. We further performed conditional analyzes to identify specific alleles or genetic variants that drive the association with PD.

**Results:** Four *HLA* types were associated with PD after correction for multiple comparisons, *HLA-DQA1*\*03:01, *HLA-DQB1*\*03:02, *HLA-DRB1*\*04:01 and *HLA-DRB1*\*04:04. Haplotype analyzes followed by amino-acid analysis and conditional analyzes suggested that the association is protective and primarily driven by three specific amino acid polymorphisms present in most *HLA-DRB1*\*04 subtypes - 11V, 13H and 33H (OR=0.87 95%CI=0.83-0.90, *p*<8.23×10^−9^ for all three variants). No other effects were present after adjustment for these amino acids.

**Conclusions:** Our results suggest that specific variants in the *HLA-DRB1* gene are associated with reduced risk of PD, providing additional evidence for the role of the immune system in PD. Although effect size is small and has no diagnostic significance, understanding the mechanism underlying this association may lead to identification of new targets for therapeutics development.

## Introduction

Although Parkinson’s disease (PD) is primarily a neurodegenerative disorder, the role of the immune system in the pathophysiology of PD is increasingly recognized based on animal and human studies.^1-3^ The immune system can be involved in the initiation of PD, as well as in its progression, and that this involvement can be peripheral and central.^3,4^

Neuropathological studies have shown evidence for microglia activation in brains of patients. However, it was initially unclear whether this activation was a part of the disease process, a consequence, or an epiphenomenon.^5^ Genetic evidence also links the immune system with PD, since genes such as *LRRK2*, the human leukocyte antigen (*HLA*) locus and possibly *BST1*, all associated with PD^6^ and have a role in the immune system.^7-9^

The *HLA* region on chromosome 6 includes genes that encode components of the major histocompatibility complex (MHC).^8^ Several genome-wide association studies (GWASs) have shown an association between the *HLA* locus and risk of PD. In the latest GWAS, an association with *HLA-DRB5* has been reported, with a potential effect of the rs112485576 single nucleotide polymorphism (SNP) on the expression of *HLA-DRB5*.^6^ Previous studies have suggested different associations with *HLA-DQA2, HLA-DQB1, HLA-DRA, HLA-DRB1, HLA-DRB5* and with haplotypes within the *HLA* region in Europeans.^10-15^

In this study, we performed the largest HLA alleles, haplotypes and amino acid analyzes in PD on 12,137 patients, 14,422 proxy patients and 351,953 controls. We further performed conditional analyzes to fine map and identify specific drivers of the association with PD in the *HLA* region.

## Methods

### Study population

This study was designed as a meta-analysis of multiple cohorts, including a total of 13,770 PD patients, 20,214 proxy-patients and 490,861 controls, as detailed in Supplementary Table 1. In brief, we included cohorts and datasets from eight independent sources: International Parkinson’s Disease Genomics Consortium (IPDGC) NeuroX dataset (dbGap phs000918.v1.p1, including datasets from multiple independent cohorts as previously described),^16^ McGill University (McGill),^17^ National Institute of Neurological Disorders and Stroke (NINDS) Genome-Wide genotyping in Parkinson’s Disease (dbGap phs000089.v4.p2),^18^ NeuroGenetics Research Consortium (NGRC) (dbGap phs000196.v3.p1),^11^ Oslo Parkinson’s Disease Study (Oslo), Parkinson’s Progression Markers Initiative (PPMI), Vance (dbGap phs000394) and PD cases and proxy-cases from the UK Biobank (UKB). Proxy-cases are first degree relatives of PD patients, thus sharing ∼50% of the patients’ genetic background and eligible to serve as proxies, as previously described.^19^ All cohorts were previously included in the most recent PD GWAS.^6^ Study protocols were approved by the relevant Institutional Review Boards and all patients signed informed consent before participating in the study.

### Pre-imputation genotype quality control

In order to include only high-quality samples and SNPs, standard quality control (QC) was performed on all datasets individually using PLINK v1.9.^20^ Standard GWAS QC was done to filter out samples and SNPs with low call rate, heterozygote outliers along with gender mismatch as previously described.^6^ SNPs deviating from Hardy-Weinberg equilibrium were removed. Only samples of European ancestry clustering with HapMap v3 using principal component analysis were included as shown in Supplementary Figure 1. In order to exclude related individuals, we examined relatedness in each dataset separately, followed by relatedness test across all datasets combined, to exclude individuals who were included in more than one dataset. All individuals with pi_hat >0.125 were excluded using GCTA v1.26.0.^21^

### UK Biobank quality control

For the analysis of the UKB data, unrelated participants of European ancestry (field 22006), with low missingness rate (field 220027) were included after exclusion of heterozygosity outliers as previously described.^6^. PD patients from the UK Biobank were included based on self-report (field 20002) or based on their International Classification of disease diagnosis code (ICD-10, code G20, field 41270). From the remaining participants, proxy-cases were defined as first degree relatives (parents or siblings, field 20112-20114) of patients with PD. Principal components were calculated using flashpca^22^ after excluding related individuals as described above. The control group was divided randomly to two groups of controls: one was included in the GWAS comparing PD patients from UKB and controls, and the second was included in the GWAS comparing proxy-cases from UKB and controls. This division was done proportionally to the size of each GWAS.

### Imputation

For SNP imputation of each dataset, we used the Michigan Imputation Server on the 1,000 Genomes Project panel (Phase 3, Version 5) using Minimac3 and SHAPEIT v2.r790. Imputed UK Biobank genotyped data v3 were downloaded in July 2019. All variants with an imputation quality (*r*^*2*^) of >0.30 were labeled as soft calls and >0.80 were labeled as hard calls. Soft calls were only used together with the hard calls for polygenic risk score calculation (see below); hard calls were used for all other analyzes.

### Association analysis of common variants on chromosome 6

Prior to determining HLA types, we performed a simple association test of all SNPs located on chromosome 6, to verify that we identified the same hit in the HLA region as previously described.^6^ For this purpose, we generated summary statistics of chromosome 6 for each dataset, and used logistic regression with an additive model adjusting for age at onset for patients and age at enrollment of controls, sex and population stratification (first 10 principal components) with PLINK v2.00a2LM (25 Oct 2019).^20^ The UK Biobank data was analyzed similarly using logistic regression adjusting for age, sex, the first 10 principal components and Townsend index to account for additional potential population stratification confounders. Finally, to harmonize effects in cases and proxy-cases, summary statistics for proxy-cases were rescaled based on genome-wide association study by proxy (GWAX) as previously described.^19^ To meta-analyze the different datasets, we performed a fixed-effect meta-analysis using METAL with an inverse-variance-based model.^23^

### HLA locus analysis

#### HLA imputation

To impute specific HLA types for each individual, we inferred two field resolution HLA alleles using HIBAG v1.22.0, a statistical method for HLA type imputation in R.^24^ HIBAG was shown to be as accurate or more accurate in Europeans compared to other types of HLA imputation tools.^25^ HIBAG provided a reference panel for Europeans (n = 2,572) with high imputation accuracy for *HLA-A, HLA-B, HLA-C*, class I genes, and *HLA-DPB1, HLA-DQA1, HLA-DQB1*, and *HLA-DRB1*, class II genes. *HLA-DRB3, HLA-DRB4* and *HLA-DRB5* imputation models were trained using HIBAG^24^ on European origin sample training set (n = 3,267) genotyped on the Illumina Infinium PsychArray-24 chip and fully sequenced at 8-digit resolution for HLA loci. These models were validated in a test set (n=886) with high accuracy (Supplementary Table 2). Imputation accuracy for European *DRB1*\*04 alleles was determined for *DRB1*\*04:01 *DRB1*\*04:02 *DRB1*\*04:03, *DRB1*\*04:04, *DRB1*\*04:05, *DRB1*\*04:07, *DRB1*\*04:08. Alleles with an imputation probability of <0.5 were defined as undetermined and individuals with two or more undetermined alleles were excluded from the analysis (Supplementary Table 1 details the numbers included for each allele in each cohort after all quality control steps). To further examine imputation accuracy, the results of the *DRB1* imputation were compared against high throughput HLA sequencing in 380 PD samples from Oslo. The combined frequency of seven different *DRB1*\*04 alleles detected in sequence data was 0.15 with the 04:01 and 04:04 alleles being the most common (Supplementary Table 3). Imputation accuracy for DRB1*04 alleles was very high at 2-digit resolution (Supplementary Table 4).

#### Statistical analysis

To examine the association of HLA alleles with PD, we used R v3.6 to perform logistic regression, adjusting for age at onset, sex and the first 10 principal components. The UK Biobank dataset was also adjusted for Townsend index. Haplotype analyzes were performed using haplo.stats in R with logistic regression as stated above. Only haplotypes with posterior probability >0.2 and carrier frequency of >1% were included in the analysis. Amino acid association analyzes were performed using HIBAG after converting P-coded alleles to amino acid sequences for exon 2, 3 of HLA class I genes and exon 2 of class II genes. Amino acid associations were tested using logistic regression as described above. A polygenic risk score (PRS) was calculated using PRSice v 2.2.11 without linkage disequilibrium (LD) clumping or P thresholding.^26^ The beta weights from the summary statistics of the 90 genome-wide significant variants in the latest PD GWAS^6^ were used in the PRS. To make sure that all possible variants were included in the PRS analysis, we also performed imputation using the Haplotype Reference Consortium panel (HRC) (Version r1.1 2016) with Minimac4 and Eagle v2.4. Ambiguous variant (rs6658353) and rs112485576 from the HLA region were excluded from the PRS calculation. To examine whether secondary hits exist in the HLA region, we adjusted for significant HLA variants, HLA alleles, HLA amino acid changes and PRS, by introducing significant findings from the first analyzes as covariates in the regression models. Statistical analyzes were only performed on alleles, haplotypes and amino acid changes with more than 1% carrier frequency. *P* value significance levels were adjusted using Bonferroni correction. Meta-analysis was performed as described above. All missing data were excluded from the analyzes.

#### Code availability

The scripts used in this analysis is available at https://github.com/gan-orlab/HLA_HIBAG/.

#### Data availability statement

Anonymized data will be shared by request from any qualified investigator.

## Results

### Meta-analysis of HLA types in Parkinson’s disease suggests a single association

After standard QC, a total of 12,137 patients, 14,422 proxy patients and 351,953 controls were included in the analysis (Supplementary Table 1). As shown in Figure 1A, our SNP-level meta-analysis validated the previous association for SNP, rs112485576, in the *HLA* locus (In the current analysis: OR=0.87, 95% CI= 0.83-0.90, p=5.00×10^−13^, in the previous meta analysis: OR=0.85, 95% CI= 0.82-0.87, p=6.96×10^−28^).^6^ No residual HLA effects were found after adjustment of this SNP (Figure 1B, C), indicating that the association of this locus was primarily driven by a single genetic risk factor. We also validated previous associations for the SNPs rs17425622, rs2395163, rs3129882, rs9275326 in the *HLA* locus (Supplementary Figure 2-5).

**Figure 1.**
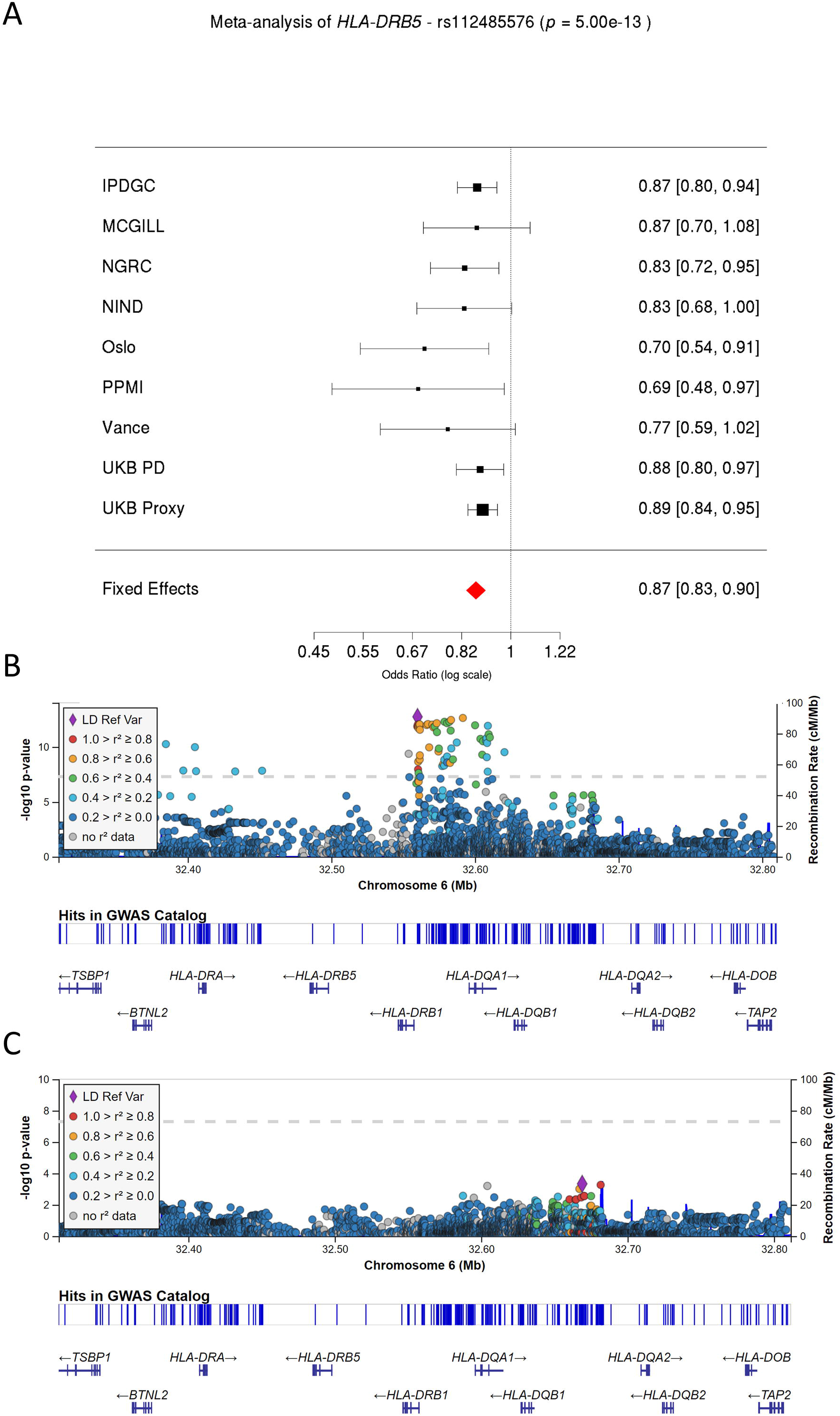
Validation of previously associated top *HLA* locus SNP (rs112485576) in our cohort. **A)** Forest plot describing the effect size and 95% confidence interval of rs112485576 for each cohort and fixed-effect meta-analysis. **B-C)** Two LocusZoom plots highlighting the significant variants before (**B**) and after (**C**) the conditional analysis on rs112485576. Dashed lines correspond to the significance threshold. Linkage disequilibrium values are shown with respect to the most significant SNP in the locus.

We next performed a meta-analysis association study of all *HLA* types with carrier frequency above 1%. After *HLA* imputation, a total of 141 different *HLA* types across 10 *HLA* loci were included (setting the Bonferroni corrected threshold for statistical significance on α=3.55×10^−4^; 0.05/141). Following these analyzes, we found four *HLA* alleles that were associated with PD (Table 1, results for other *HLA* alleles are detailed in Supplementary Table 5): *HLA-DQA1*\*03:01, *HLA-DQB1*\*03:02, *HLA-DRB1*\*04:01, *HLA-DRB1*\*04:04. These four alleles are all located within a small genomic segment and have similar odds ratio ranging between 0.84-0.89. Three of the four alleles have similar carrier frequencies, indicating that they could be part of the same haplotypes, with the fourth potentially representing a sub-haplotype (Table 1).

**Table 1.**
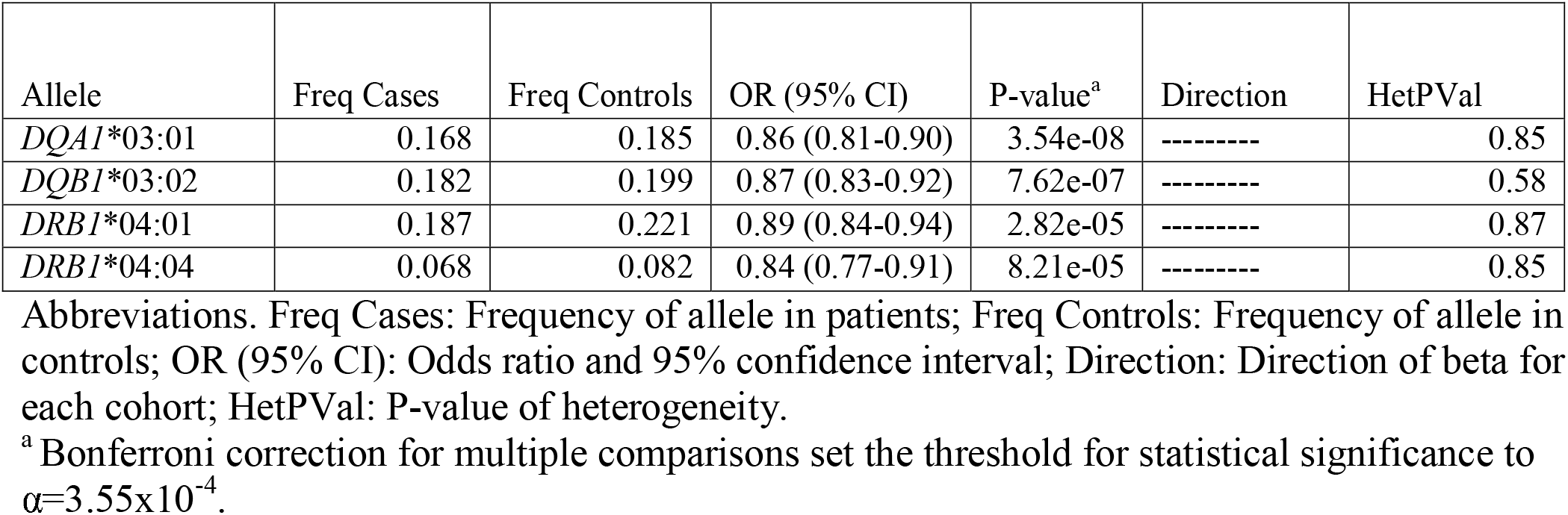
Meta-analyzes of HLA alleles association.

### HLA haplotype analysis

For haplotype analysis, we allowed for up to three genes to be included in each haplotype, since including more than three genes generated multiple haplotypes with low allele frequency that could not be analyzed at the current sample size. A total of 84 different HLA haplotypes (Supplementary Table 6) with allele frequency >1% were identified, setting the cut-off Bonferroni corrected p value for statistical significance at α=5.95×10^−4^. Three different HLA haplotypes were associated with PD after correction for multiple comparisons: *DQA1\**03:01*∼DQB1\**03:02, *DRB1\**04:01*∼DQA1\**03:03 and *DRB1\**04:04*∼DQA1\**03:01. Upon further examination, this association was found to be driven by several well-known sub-haplotypes, *DRB1*\*04:04*∼DQA1*\*03:01*∼DQB1*\*03:02 and *DRB1*\*04:01*∼DQA1*\*03:01/3*∼DQB1*\*03:01/2 (Supplementary Table 6). Because both *DQA1*\*03:01 and *DQA1*\*03:03 as well as *DQB1*\*03:01 and *DQB1*\*03:03 are present within the extended *DRB1*\*04:01 haplotype, it is likely that these associations are driven by *DRB1*.

### Meta-analysis of the association of HLA amino acid changes with Parkinson’s disease

To further identify the specific source of the association in the *HLA* locus, we performed an analysis of 636 amino acid changes in the HLA genes, setting the cut-off Bonferroni corrected p value for statistical significance at α=7.86×10^−5^. Ten amino acid changes were significantly associated with reduced risk of PD (Supplementary Table 7). The top three associated variants are linked amino-acids 11V, 13H and 33H (Table 3) present in all *DRB1*\*04 subtypes, complementing the HLA haplotype analysis. Four other variants, 26S, 47Q, 56R and 76V, in the *DQA1* gene, are in perfect LD with each other (r^2^=1, D’=1, Supplementary Table 7) and in partial LD (r^2^=0.38, D’=0.85) with the *DRB1* variants. The association of these *DQA1* variants is weaker than the *DRB1* variants in terms of both effect size and statistical association (Supplementary Table 7). Three other variants, 71T, 74E, and 75L, are in the *DQB1* gene, are also in perfect LD with each other (r^2^=1, D’=1, Supplementary Table 7) and in partial LD (r^2^=0.16, D’=0.99) with *DRB1* 13H and 33H.

**Table 2.**
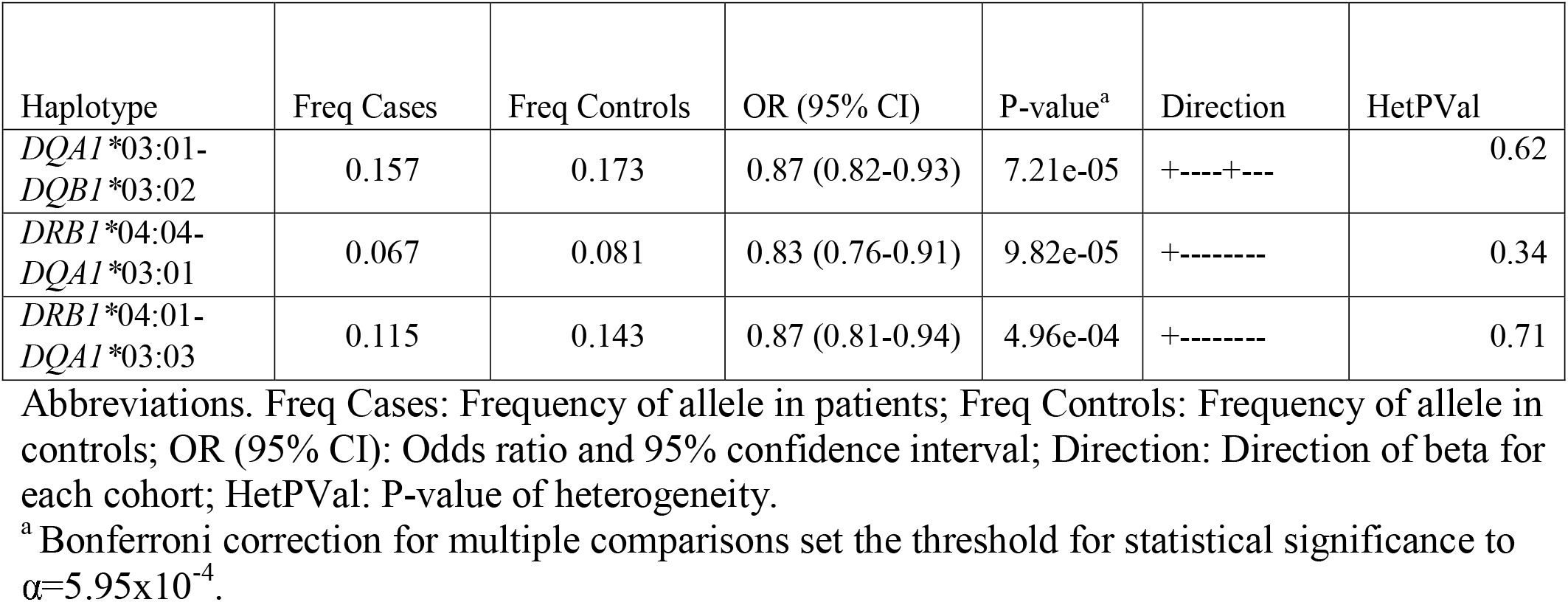
Meta-analyzes of HLA haplotype association.

**Table 3.**
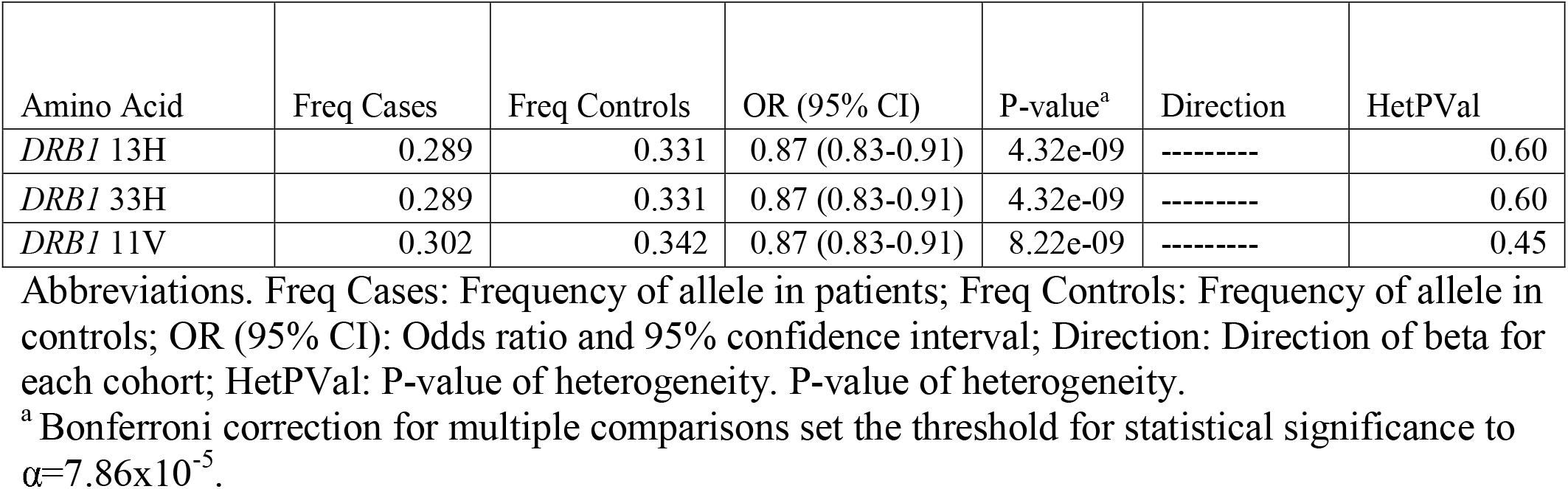
Meta-analyzes of HLA amino acid changes association.

### Conditional analyzes confirm that *DRB1*\*04 amino acid variants likely drive the association of the *HLA* locus with PD

To further determine the specific genes or variants that drive these associations, we performed a set of conditional analyzes, and re-analyzed the allele types, haplotypes and amino acid associations with PD. We conditioned the HLA type regression model on the following: rs112485576 (the top GWAS hit in the *HLA* locus), *DQA1*\*03:01, *DQA1*\*03:03 and the *DRB1* variant 13H. We have also adjusted for the PD PRS, to examine a potential polygenic effect (Supplementary Tables 5, 7). While the adjustment for the *DRB1* variant 13H completely eliminated the associations in the *DQA1* gene, adjustment for *DQA1*\*03:01 and *DQA1*\*03:03 did not completely eliminate the association of the *DRB1* gene (Supplementary Table 5), again supporting this gene and these specific amino acids (11V, 13H and 33H, Table 3) as the drivers of the association in the *HLA* locus. Adjustment for PRS did not change the results. It is also worth noting that the *DRB1* variants 11V (r^2^=0.96, D’=0.99), 13H (r^2^=0.99, D’=0.99), and 33H (r^2^=0.99, D’=0.99) are in LD with rs112485576.

## Discussion

In the current study, we performed a thorough analysis of the *HLA* region and examined its association with PD in the European population using a total of 12,137 patients, 14,422 proxy patients and 351,953 controls. Following a series of regression models and conditional analyzes, our results indicate that the drivers of the association in the *HLA* region are three amino acid changes specific of *HLA-DRB1*\*04 subtypes, 11V, 13H and 33H (Figure 2). Two of these amino acid changes, 13H and 33H are in perfect LD, and 11V is in very strong LD with the other two variants. This study agrees with a smaller HLA sequencing study^12^ in 1,597 PD cases and 1,606 controls which also observed a protective effect of *DRB1*\*04 and the same amino acids, although it also reported additional associations with *DRB1*\*01:01 and *DRB1*10:01* which were not confirmed in the current study. Interestingly, the V-H-H motif at position 11V, 13H and 33H are central to the DRB1*04:01 heterodimer and contribute to peptide binding, notably through pocket P6^27^ (Figure 2).

**Figure 2.**
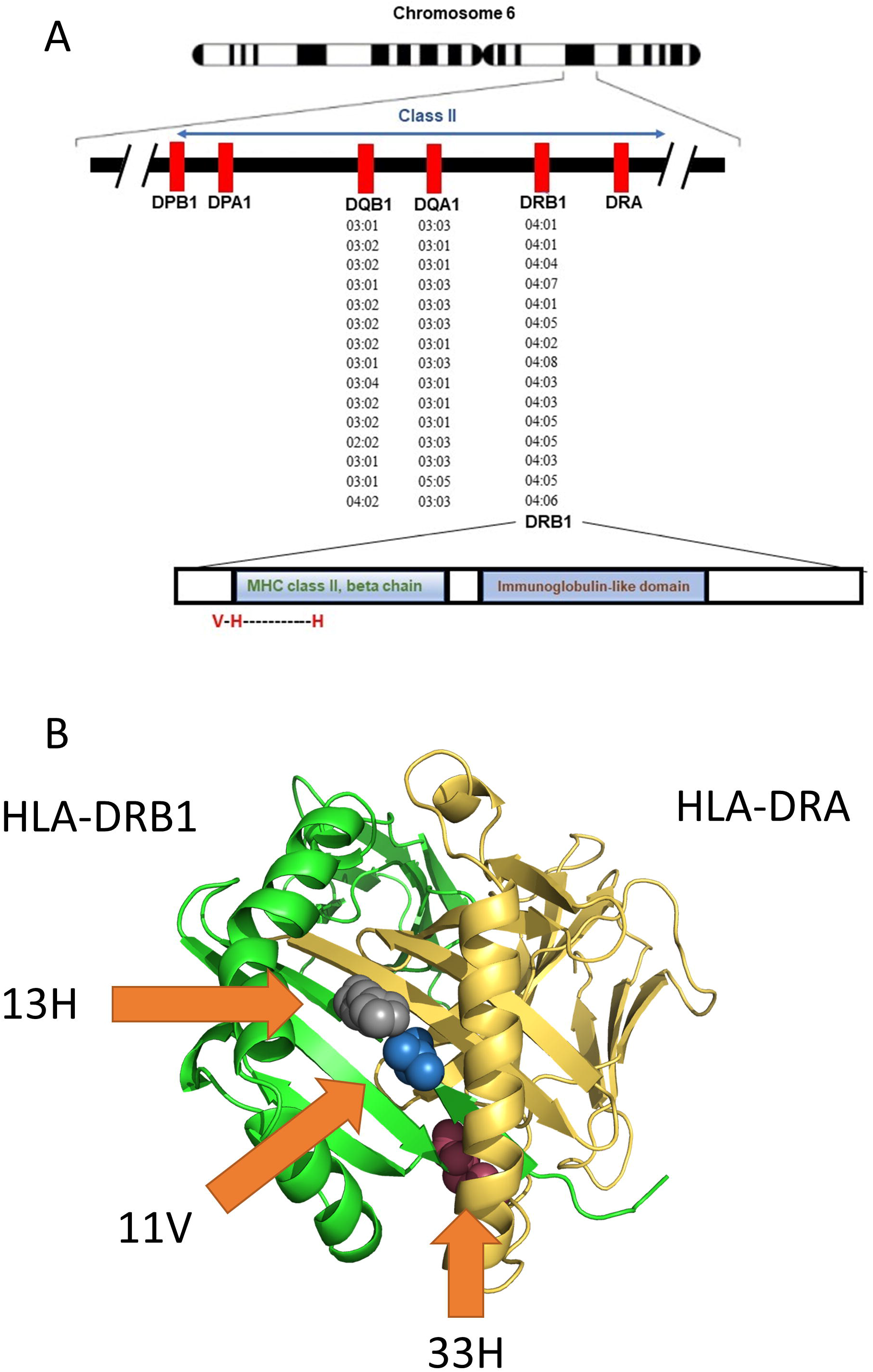
Association of the *HLA-DRB1* alleles and location of associated amino acids. **A)** The location of the HLA locus, alleles and amino acids associated with Parkinson’s disease in the current study. **B)** 3D model of HLA-DRB1 – HLA-DRA and the location of the 11V, 13H and 33H amino acids associated with PD (highlighted by arrows). The model was generated with PyMol v. 2.4.1 (pdb 4is6).

Previous studies on the *HLA* genomic region in PD have reported associations of different genes and HLA types with PD, including *HLA-DQA2, HLA-DQB1, HLA-DRA, HLA-DRB1* and *HLA-DRB5*.^10-15^ The suggested association with *HLA-DRB5* reported in the most recent PD GWAS is based on an expression quantitative trait locus (eQTL) analysis, as the top associated SNP in this region, rs112485576, was also associated with differential expression of *HLA-DRB*.^6^ A previous study of 2,000 PD patients and 1,986 controls has implicated a non-coding variant (rs3129882) within *HLA-DRA* as driving the association with PD and suggested that this variant affects the expression of *HLA-DR* and *HLA-DQ* genes.^11^ Similarly, another study suggested that the same variant in *HLA-DRA* (rs3129882) is associated with differential expression of MHC-II on immune cells.^28^ While our study does not rule out this possibility, since the main variants driving the association are amino acid changes in *DRB1*\*04 that will affect epitope binding ability, it is likely that the effect on PD risk is through these variants and not due to modified expression. Additional functional studies will be required to study this hypothesis.

The current study adds further support to the hypothesis suggesting an involvement of the peripheral and central immune system in PD. On top of the *HLA* locus, several other genes with potential roles in the immune system, including *LRRK2* and potentially *BST1*,^7, 9^ have been implicated in PD.^6^ In the periphery, there are notable changes in the immune system of PD patients compared to controls, as peripheral monocytes have differential expression of immune related proteins and markers.^3^ Whether these changes are drivers of the disease or a result of the disease is still undetermined, but accumulating evidence suggest that they can be part of the pathogenic process of PD. In the central nervous system, pathological studies suggest that microglia cells may have a central role in PD.^29^ Microgliosis is a prominent pathological finding in post-mortem brains of PD patients, and evidence suggest that microglia activation occurs early in the disease process and may be involved in the pathogenesis of PD.^3^ The specific contribution of *HLA* to these processes is still unclear and needs to be further studied.

One intriguing possibility that may directly involve *HLA* with PD is the potential interaction of HLA-DRB1*04 with α-synuclein, notably an epitope surrounding p.S129. Recent data has shown that α-synuclein fragments can bind MHC and increase T cell reactivity.^30^ This activity is proinflammatory, involves both CD4 and CD8 cells and may occur before the onset of motor symptoms,^30, 31^ suggesting an involvement of inflammation in early PD pathogenesis. Studying specific α-synuclein fragments has suggested that two major regions of α-synuclein may be associated with increased T cell reactivity in PD, with preferential CD4 activity: an N-terminal region involving amino acid p.Y39 and a C-terminal region surrounding amino acid p.S129, two important residues undergoing phosphorylation.^30^ The phosphorylation of p.S129 is particularly interesting as it is well known in promoting aggregation.^32, 33^ Further analysis focused on the p.Y39 region suggested an association with α-synuclein-specific p.Y39 T cell responses and HLA DRB1*15:01 and DRB5*01:01 presentation. This association was abolished by phosphorylation, which reduced binding of p.Y39-phosphorylated α-synuclein.^30, 31^ Other experiments by these authors have also suggested CD8+ T cells responses mediated by HLA-A11*01 presentation of epitopes in the same N-terminal region of α-synuclein. However, in the current study we could not confirm an association of these HLA types with PD.

More interestingly in the context of our work, increased CD4+ T cell response to both p.S129 phosphorylated and unphosphorylated α-synuclein was also demonstrated, suggesting involvement of DQB1*05:01 and DQB1*04:02, as these alleles strongly bound these α-synuclein epitopes.^30^ These HLA alleles, however, are not associated with risk of PD in the current study. However, the authors reported in the supplementary data that DRB1*04:01 was also a selective and strong binder of the same α-synuclein epitope with p.S129, but only when the epitope was unphosphorylated.^30^ Notably, no other DRB1 alleles that were assayed in this study^30^ had increased binding affinity to α-synuclein epitope with p.S129, except for the DRB1*04:01 allele that was a strong binder only when unphosphorylated. Binding register analysis using Immune Epitope Database (IEDB) MHC-II Binding Prediction (http://tools.iedb.org/mhcii/) suggests that this epitope binds a 9 amino acid AYEMPSEEG core, with p.S129 at P6 position, a position postulated to be important based on our HLA-DR amino acid analysis presented above. As CD4+ T cell responses are generally stronger when epitopes are presented by HLA-DR versus HLA-DQ,^34^ HLA-DRB1*04 responses to the p.S129 unphosphorylated form of α-synuclein could be dominant in individuals with HLA-DRB1*04, explaining the protective effect of this HLA subtype in PD. Additional experiments will be needed to further explore this hypothesis.

Our study has several limitations. First, this study was performed on European populations, and the results may be limited to this population only. Additional studies in other populations are required. Several studies on *HLA* types and PD have been performed in Asian populations,^35-38^and the GWAS risk variant rs112485576 has a similar OR (0.85) in the largest Asian GWAS to date,^39^ yet larger studies are required, as well as studies in other populations. An additional potential limitation of our study is its use of imputation rather than fully sequenced HLA types. Given the very high performance of the imputation tool when compared to full sequencing (Supplementary Table 2), the potential effect of imputation inaccuracies is likely small and should be diluted in our large sample size. In addition, we cannot rule out that other, rarer HLA types that were not included in the current analysis may also have a role in PD. An additional limitation of the current study is that by adjusting for sex we eliminate potential sex-specific effects. It is possible that specific HLA types are relevant in one sex more or less than the other, and this should be studied in larger, sex-stratified cohorts.

To conclude, our results suggest a role for the *HLA-DRB1* gene in susceptibility for PD, and provide further evidence for the importance of the immune system in PD. Since the effect is small, it does not merit routine HLA typing in PD, but understanding the mechanism underlying this association may lead to better understanding of PD in general and offer new targets for future immune-related treatment.

## Data Availability

Code used for the analysis can be found at https://github.com/gan-orlab/. Summary statistics are included in the supplementary files. All other data is available upon request.

## Glossary

eQTL: expression quantitative trait locus
GWAS: genome-wide association study
GWAX: genome-wide association study by proxy
HLA: human leukocyte antigen
LD: linkage disequilibrium
MHC: major histocompatibility complex
PD: Parkinson’s disease
PRS: polygenic risk score
QC: quality control
SNP: single nucleotide polymorphism

## Acknowledgment

We would like to thank the participants in the different cohorts. The access to part of the participants for this research has been made possible thanks to the Quebec Parkinson’s Network (http://rpq-qpn.ca/en/). in collaboration with Parkinson Quebec, and by the Young Investigator Award by Parkinson Canada.

## Appendix 1: Authors

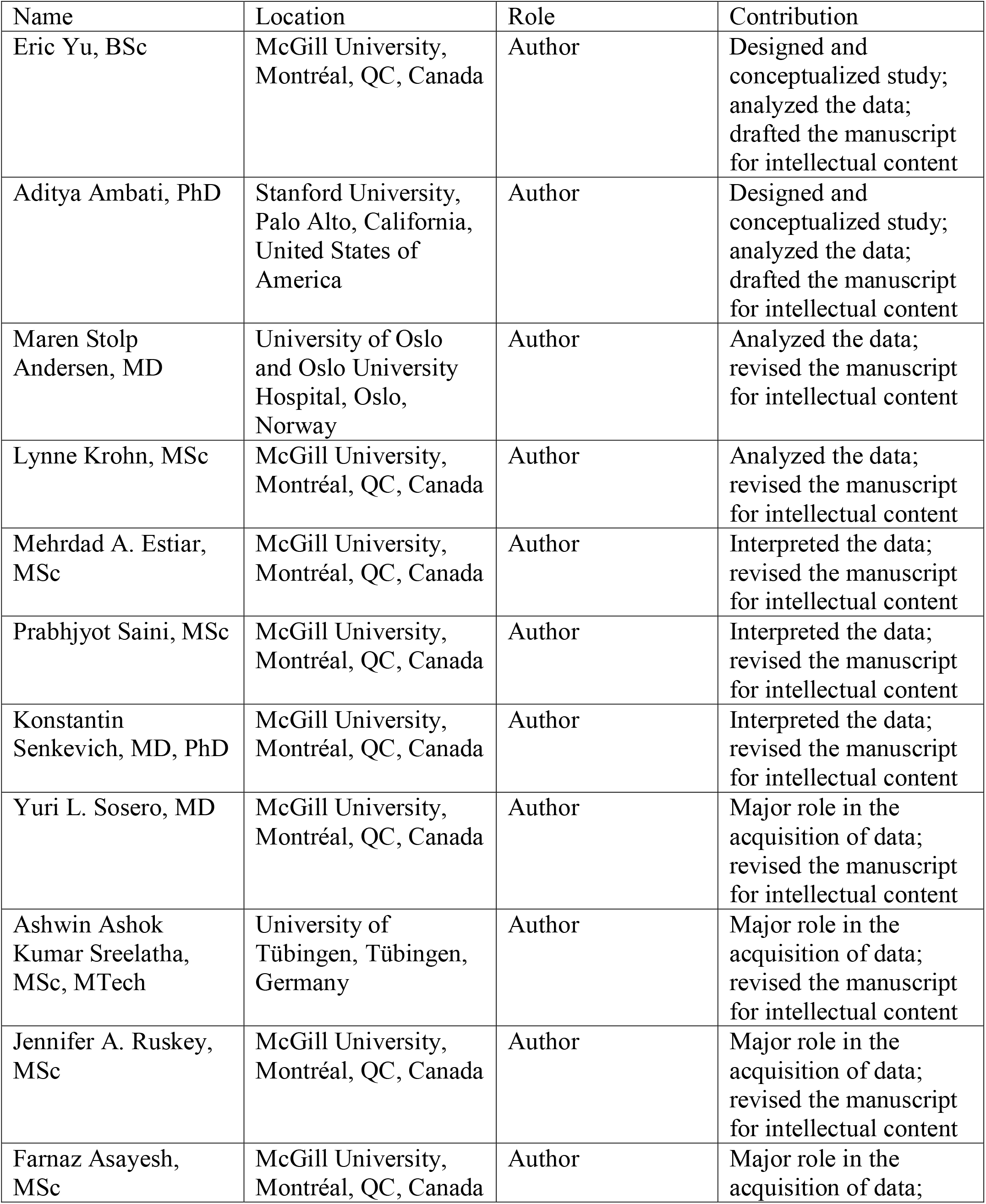
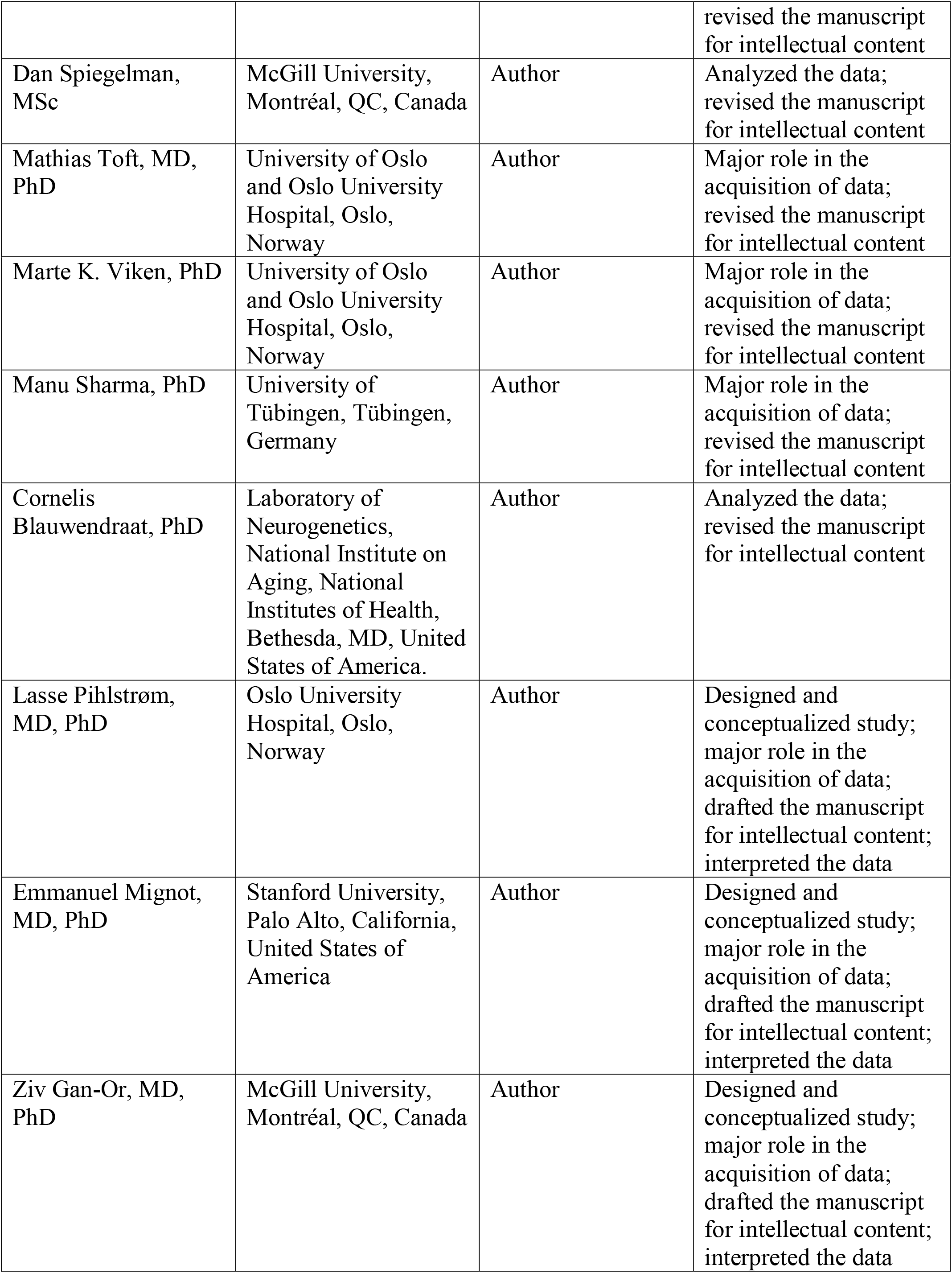

## References

1. Sanchez-Guajardo V, Barnum CJ, Tansey MG, Romero-Ramos M. Neuroimmunological processes in Parkinson’s disease and their relation to α-synuclein: microglia as the referee between neuronal processes and peripheral immunity. ASN neuro 2013;5:AN20120066.

2. Wang Q, Liu Y, Zhou J. Neuroinflammation in Parkinson’s disease and its potential as therapeutic target. Translational Neurodegeneration 2015;4:19.

3. Tansey MG, Romero-Ramos M. Immune system responses in Parkinson’s disease: Early and dynamic. Eur J Neurosci 2019;49:364–383.

4. Devos D, Lebouvier T, Lardeux B, et al. Colonic inflammation in Parkinson’s disease. Neurobiology of disease 2013;50:42–48.

5. Zhang W, Wang T, Pei Z, et al. Aggregated α-synuclein activates microglia: a process leading to disease progression in Parkinson’s disease. The FASEB Journal 2005;19:533–542.

6. Nalls MA, Blauwendraat C, Vallerga CL, et al. Identification of novel risk loci, causal insights, and heritable risk for Parkinson’s disease: a meta-analysis of genome-wide association studies. Lancet Neurol 2019;18:1091–1102.

7. Malavasi F, Deaglio S, Ferrero E, et al. CD38 and CD157 as receptors of the immune system: a bridge between innate and adaptive immunity. Mol Med 2006;12:334–341.

8. Shiina T, Hosomichi K, Inoko H, Kulski JK. The HLA genomic loci map: expression, interaction, diversity and disease. J Hum Genet 2009;54:15–39.

9. Wallings RL, Herrick MK, Tansey MG. LRRK2 at the Interface Between Peripheral and Central Immune Function in Parkinson’s. Front Neurosci 2020;14:443.

10. Ahmed I, Tamouza R, Delord M, et al. Association between Parkinson’s disease and the HLA-DRB1 locus. Mov Disord 2012;27:1104–1110.

11. Hamza TH, Zabetian CP, Tenesa A, et al. Common genetic variation in the HLA region is associated with late-onset sporadic Parkinson’s disease. Nat Genet 2010;42:781–785.

12. Hollenbach JA, Norman PJ, Creary LE, et al. A specific amino acid motif of HLA-DRB1 mediates risk and interacts with smoking history in Parkinson’s disease. Proc Natl Acad Sci U S A 2019;116:7419–7424.

13. Wissemann WT, Hill-Burns EM, Zabetian CP, et al. Association of Parkinson disease with structural and regulatory variants in the HLA region. The American Journal of Human Genetics 2013;93:984–993.

14. Bandres-Ciga S, Ahmed S, Sabir MS, et al. The Genetic Architecture of Parkinson Disease in Spain: Characterizing Population-Specific Risk, Differential Haplotype Structures, and Providing Etiologic Insight. Mov Disord 2019;34:1851–1863.

15. Hill-Burns EM, Factor SA, Zabetian CP, Thomson G, Payami H. Evidence for more than one Parkinson’s disease-associated variant within the HLA region. PLoS One 2011;6:e27109.

16. Nalls MA, Pankratz N, Lill CM, et al. Large-scale meta-analysis of genome-wide association data identifies six new risk loci for Parkinson’s disease. Nat Genet 2014;46:989–993.

17. Gan-Or Z, Rao T, Leveille E, et al. The Quebec Parkinson Network: A Researcher-Patient Matching Platform and Multimodal Biorepository. 2020:1–13.

18. Simón-Sánchez J, Schulte C, Bras JM, et al. Genome-wide association study reveals genetic risk underlying Parkinson’s disease. Nat Genet 2009;41:1308–1312.

19. Liu JZ, Erlich Y, Pickrell JK. Case-control association mapping by proxy using family history of disease. Nat Genet 2017;49:325–331.

20. Chang CC, Chow CC, Tellier LC, Vattikuti S, Purcell SM, Lee JJ. Second-generation PLINK: rising to the challenge of larger and richer datasets. GigaScience 2015;4.

21. Yang J, Lee SH, Goddard ME, Visscher PM. GCTA: a tool for genome-wide complex trait analysis. Am J Hum Genet 2011;88:76–82.

22. Abraham G, Inouye M. Fast principal component analysis of large-scale genome-wide data. PLoS One 2014;9:e93766.

23. Willer CJ, Li Y, Abecasis GR. METAL: fast and efficient meta-analysis of genomewide association scans. Bioinformatics 2010;26:2190–2191.

24. Zheng X, Shen J, Cox C, et al. HIBAG—HLA genotype imputation with attribute bagging. The pharmacogenomics journal 2014;14:192–200.

25. Karnes JH, Shaffer CM, Bastarache L, et al. Comparison of HLA allelic imputation programs. PLoS One 2017;12:e0172444.

26. Choi SW, O’Reilly PF. PRSice-2: Polygenic Risk Score software for biobank-scale data. Gigascience 2019;8:giz082.

27. Kamoun M, McCullough KP, Maiers M, et al. HLA Amino Acid Polymorphisms and Kidney Allograft Survival. Transplantation 2017;101:e170–e177.

28. Kannarkat GT, Cook DA, Lee JK, et al. Common Genetic Variant Association with Altered HLA Expression, Synergy with Pyrethroid Exposure, and Risk for Parkinson’s Disease: An Observational and Case-Control Study. NPJ Parkinsons Dis 2015;1.

29. Levesque S, Wilson B, Gregoria V, et al. Reactive microgliosis: extracellular micro-calpain and microglia-mediated dopaminergic neurotoxicity. Brain 2010;133:808–821.

30. Sulzer D, Alcalay RN, Garretti F, et al. T cells from patients with Parkinson’s disease recognize α-synuclein peptides. Nature 2017;546:656–661.

31. Lindestam Arlehamn CS, Dhanwani R, Pham J, et al. α-Synuclein-specific T cell reactivity is associated with preclinical and early Parkinson’s disease. Nat Commun 2020;11:1875.

32. Fayyad M, Erskine D, Majbour NK, et al. Investigating the presence of doubly phosphorylated α-synuclein at tyrosine 125 and serine 129 in idiopathic Lewy body diseases. Brain Pathol 2020;30:831–843.

33. Li XY, Yang W, Li X, et al. Phosphorylated Alpha-Synuclein in Red Blood Cells as a Potential Diagnostic Biomarker for Multiple System Atrophy: A Pilot Study. Parkinsons Dis 2020;2020:8740419.

34. Grifoni A, Moore E, Voic H, et al. Characterization of Magnitude and Antigen Specificity of HLA-DP, DQ, and DRB3/4/5 Restricted DENV-Specific CD4+ T Cell Responses. Front Immunol 2019;10:1568.

35. Chang KH, Wu YR, Chen YC, Fung HC, Chen CM. Association of genetic variants within HLA-DR region with Parkinson’s disease in Taiwan. Neurobiol Aging 2020;87:140 e113–140 e118.

36. Ma ZG, Liu TW, Bo YL. HLA-DRA rs3129882 A/G polymorphism was not a risk factor for Parkinson’s disease in Chinese-based populations: a meta-analysis. Int J Neurosci 2015;125:241–246.

37. Sun C, Wei L, Luo F, et al. HLA-DRB1 alleles are associated with the susceptibility to sporadic Parkinson’s disease in Chinese Han population. PLoS One 2012;7:e48594.

38. Zhao Y, Gopalai AA, Ahmad-Annuar A, et al. Association of HLA locus variant in Parkinson’s disease. Clin Genet 2013;84:501–504.

39. Foo JN, Chew EGY, Chung SJ, et al. Identification of Risk Loci for Parkinson Disease in Asians and Comparison of Risk Between Asians and Europeans: A Genome-Wide Association Study. JAMA Neurol 2020;77:746–754.

