## Supplementary figures and images for "Fine mapping of the *HLA* locus in Parkinson’s disease in Europeans"

### Supplemental Figure 1

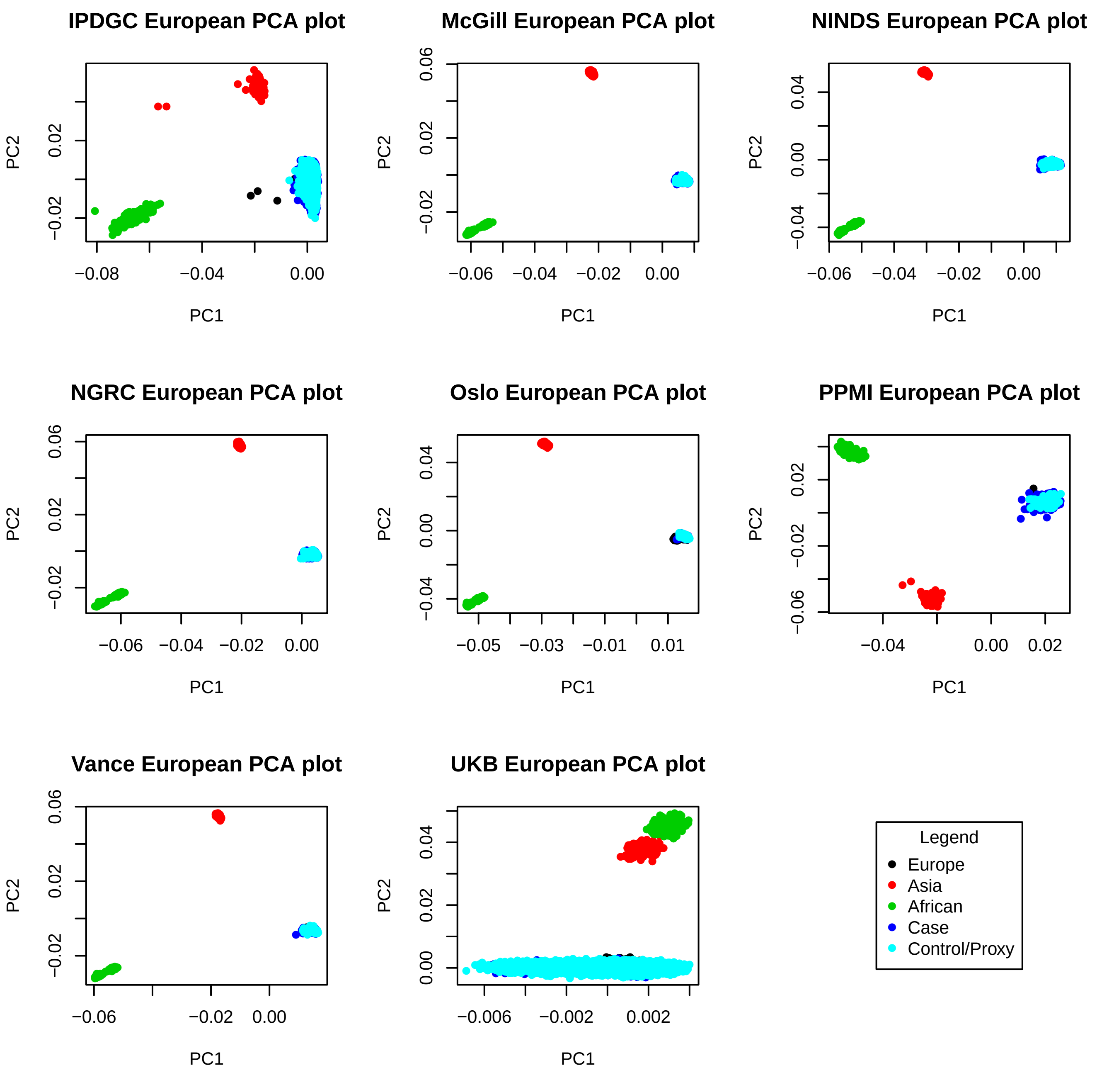

### Supplemental Figure 2

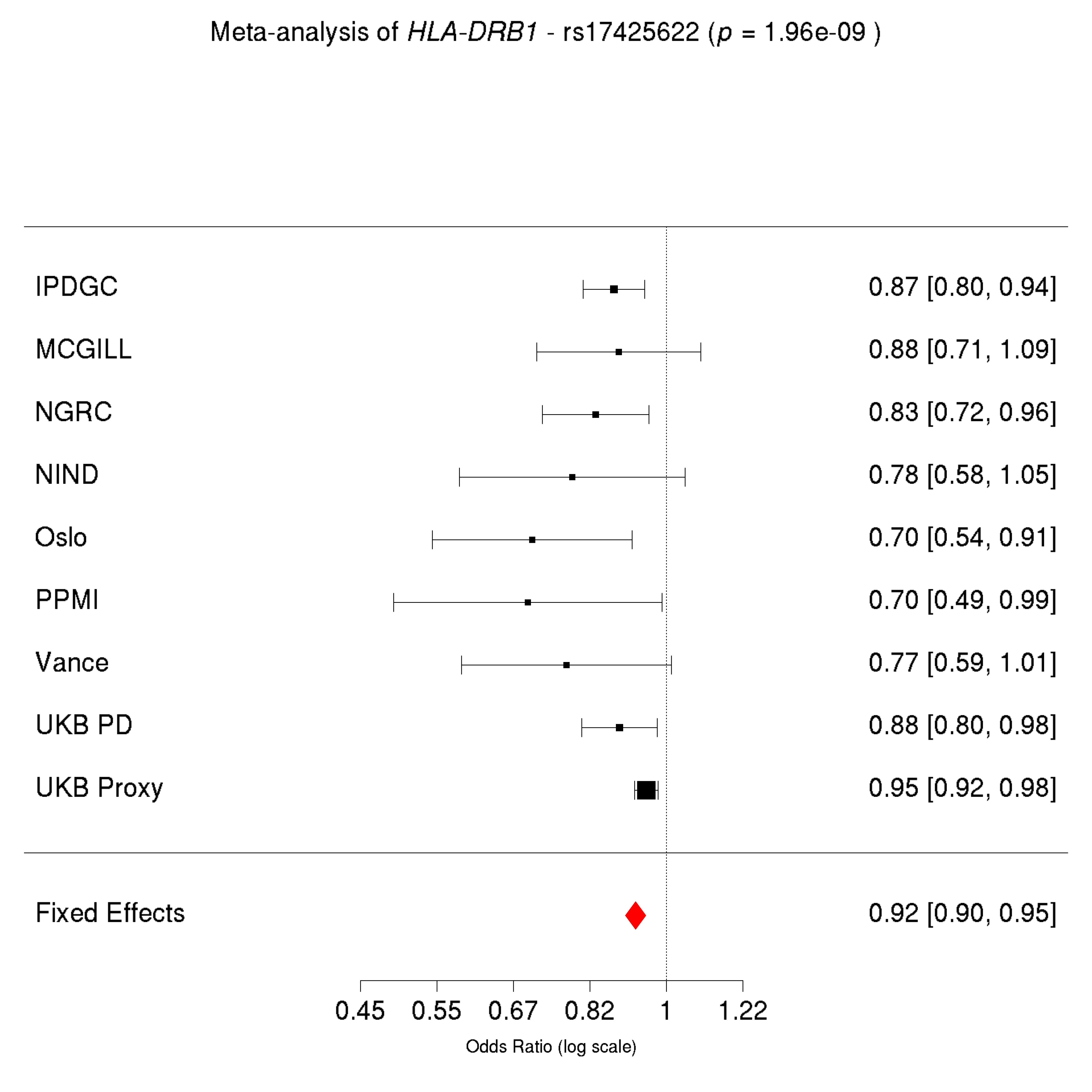

### Supplemental Figure 3

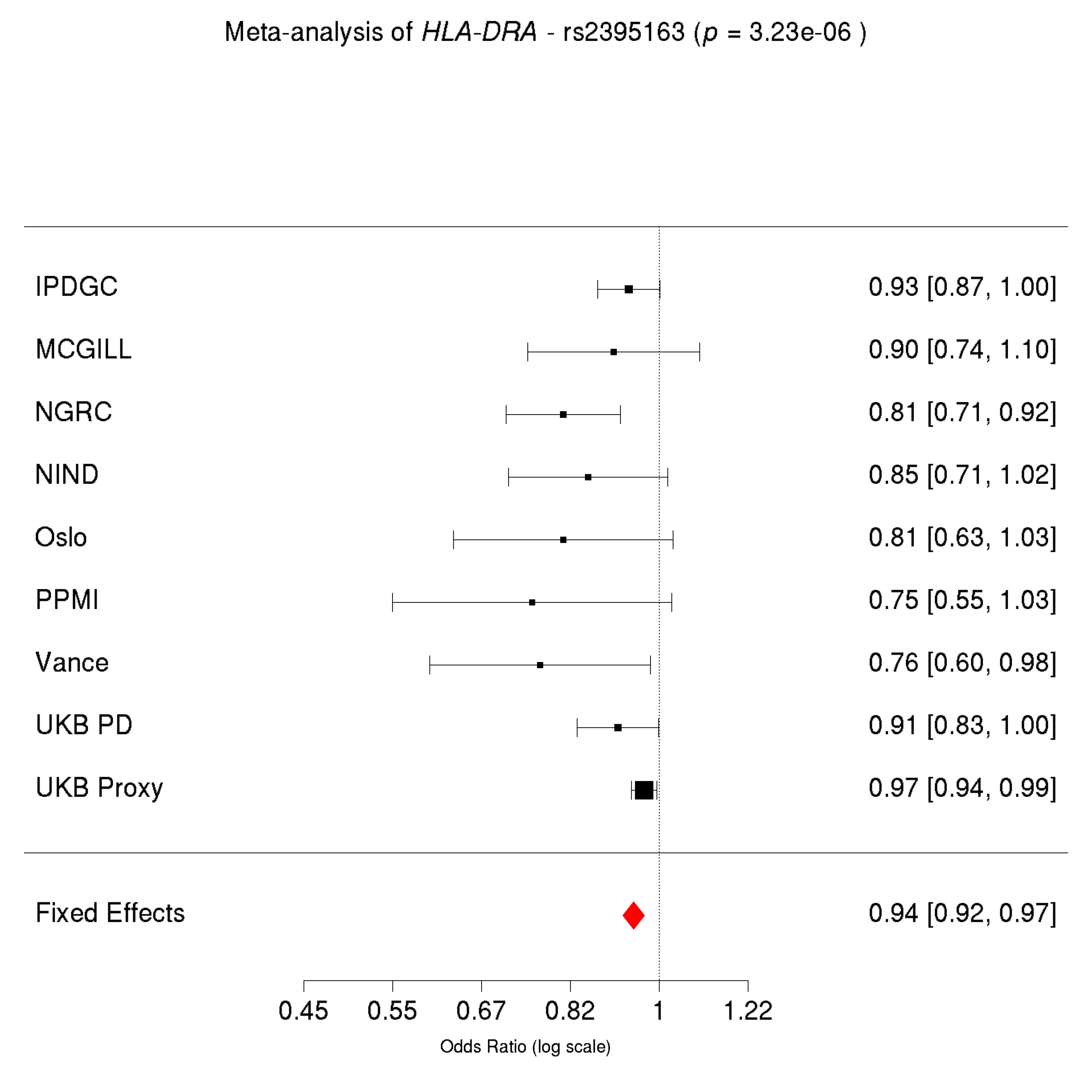

### Supplemental Figure 4

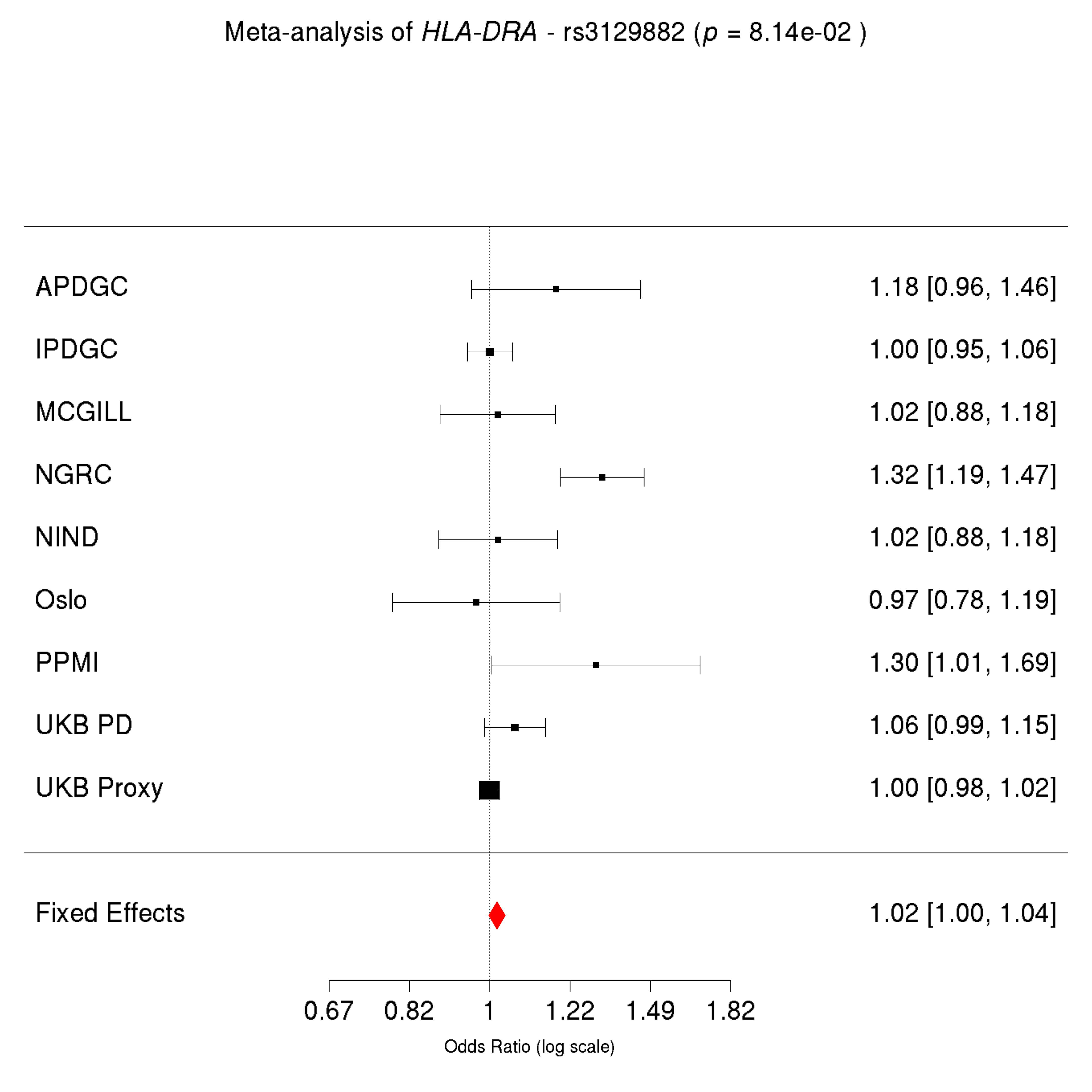

### Supplemental Figure 5

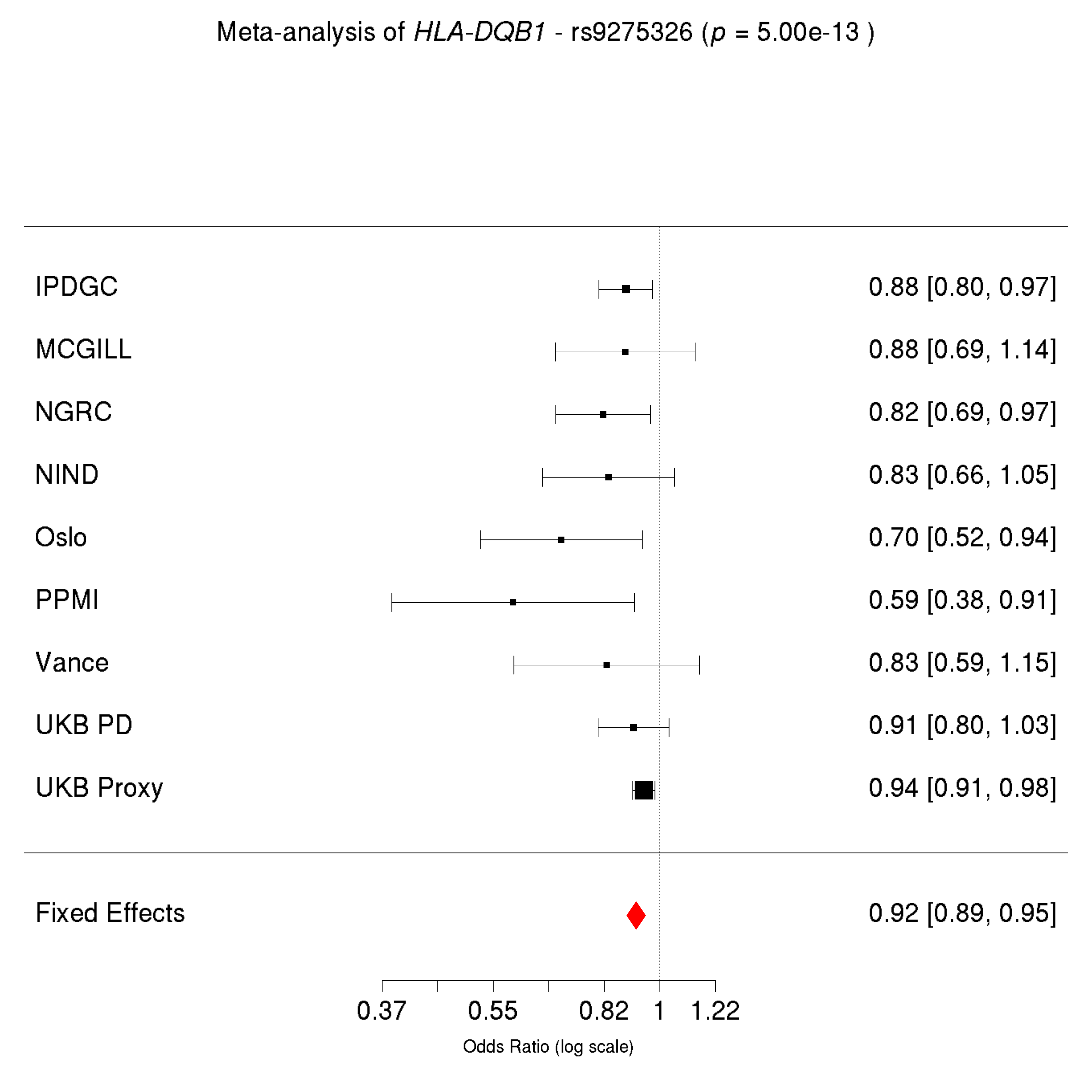
